# High Liver Fat Associates with Higher Risk of Developing Symptomatic COVID-19 Infection - Initial UK Biobank Observations

**DOI:** 10.1101/2020.06.04.20122457

**Authors:** A Roca-Fernández, A Dennis, R Nicolls, J McGonigle, M Kelly, R Banerjee

## Abstract

**Background:** A high proportion of COVID-19 patients develop acute liver dysfunction. Early research has suggested that pre-existing fatty liver disease may be a significant risk factor for hospitalisation. Liver fat, in particular, is a modifiable parameter and can be a target for public health policy and individual patient plans. In this study we aimed to assess pre-existing liver disease as a risk factor for developing symptomatic COVID-19.

**Methods:** From 502,506 participants from the UK Biobank, 42,146 underwent MRI (aged 45–82), and had measures of liver fat, liver fibroinflammatory disease and liver iron. Patients were censored on May 28th to determine how many had tested for COVID-19 with symptomatic disease. UK testing was restricted to those with symptoms in hospital. COVID-19 symptoms included fever, dry cough, sore throat, diarrhoea and fatigue. Univariate analysis was performed on liver phenotypic biomarkers to determine if these variables increased risk of symptomatic COVID-19, and compared to previously described risk factors associated with severe COVID-19, including to age, ethnicity, gender and obesity,

**Findings:** Increased liver fat was associated with a higher risk for symptomatic confirmed COVID-19 in this population in univariate analysis(OR:1.85, p = 0.03). In obese participants, only those with concomitant fatty liver(≥10%) were at increased risk(OR:2.96, p = 0.02), with those having normal liver fat (< 5%) showing no increased risk(OR:0.36, p = 0.09).

**Conclusions:** UK Biobank data demonstrated an association between pre-existing liver disease and obesity with severe COVID-19, with higher proportions of liver fat in obese individuals a likely risk factor for symptomatic disease and severity.

Public policy measures to protect patients with liver disease who may have almost double the risk of the general population should be considered, especially as dietary and pharmacological strategies to reduce body weight and liver fat already exist.

**Funding:** University of Oxford, Innovate UK, UK Biobank. Authors are employees of Perspectum Ltd.

## INTRODUCTION

Coronavirus disease (COVID-19) is an infectious disease caused by the recently discovered SARS-CoV-2 coronavirus. Risk factors leading to severe infection have been identified in previous studies around the world (1–3) including increased age, male sex, non-Caucasian ethnicity and the presence of pre-existing co-morbidities, in particular cardiovascular and metabolic conditions, particularly those conditions involving liver function.

The majority of patients experiencing COVID-19 developed abnormal liver function(4), with studies reporting abnormal alanine transaminase (ALT), aspartate aminotransferase (AST), albumin levels, and increased gamma-glutamyl transferase (GGT) serum in severe patients (6). The development of liver injury was more common in severe COVID-19, with liver injury in mild disease often transient and resolving without treatment.

The mechanistic link between COVID-19 and the liver may relate to the expression of the ACE2 protein, through which SARS-CoV-2 is believed to enter cells(7) in cholangiocytes (8). However, recent studies have also shown a hepatocellular profile of damage rather than cholestasis (9). Alternative hypotheses suggest that liver injury in severe cases is being caused by an immune response triggered by the infection (6) or due to a drug-induced liver injury from the antivirals, antibiotics and steroids widely being used for treating COVID-19 (10).

Traditionally, characterisation of chronic liver disease has relied on invasive liver biopsy to evaluate the pathological hallmarks of disease: fibrosis, inflammation, and steatosis(11). More recently, scalable, non-invasive alternatives with precise and accurate quantitative measurements have been developed using MRI biomarkers of liver fat (also known as Proton Density Fat Fraction, PDFF, and fibroinflammation, cT1). Multiparametric MRI measuring liver fat and fibroinflammation has demonstrated value in characterising liver disease(12,13) and predicting clinical outcomes(14). This has allowed for determination of the current prevalence of fatty liver disease in the UK adult population, along with genetic associations with liver fat and fibroinflammation(15). The UK Biobank has recently released COVID-19 data on testing status for a subset of participants, thus offering a unique resource for investigating the associations between demographic, phenotypic and genotypic factors with COVID-19 disease severity and related healthcare outcomes.

The aim of this study was to test the hypothesis that liver disease, and specifically liver fat accumulation, is a risk factor for developing symptomatic COVID-19.

## METHODS

42,146 participants were extracted from the UK Biobank (UKB) imaging study with complete liver imaging to measure fat and fibroinflammatory disease. Of these, 287 had a COVID-19 test result available with 79 testing positive and 208 testing negatives (Figure 1).

**Figure 1:**
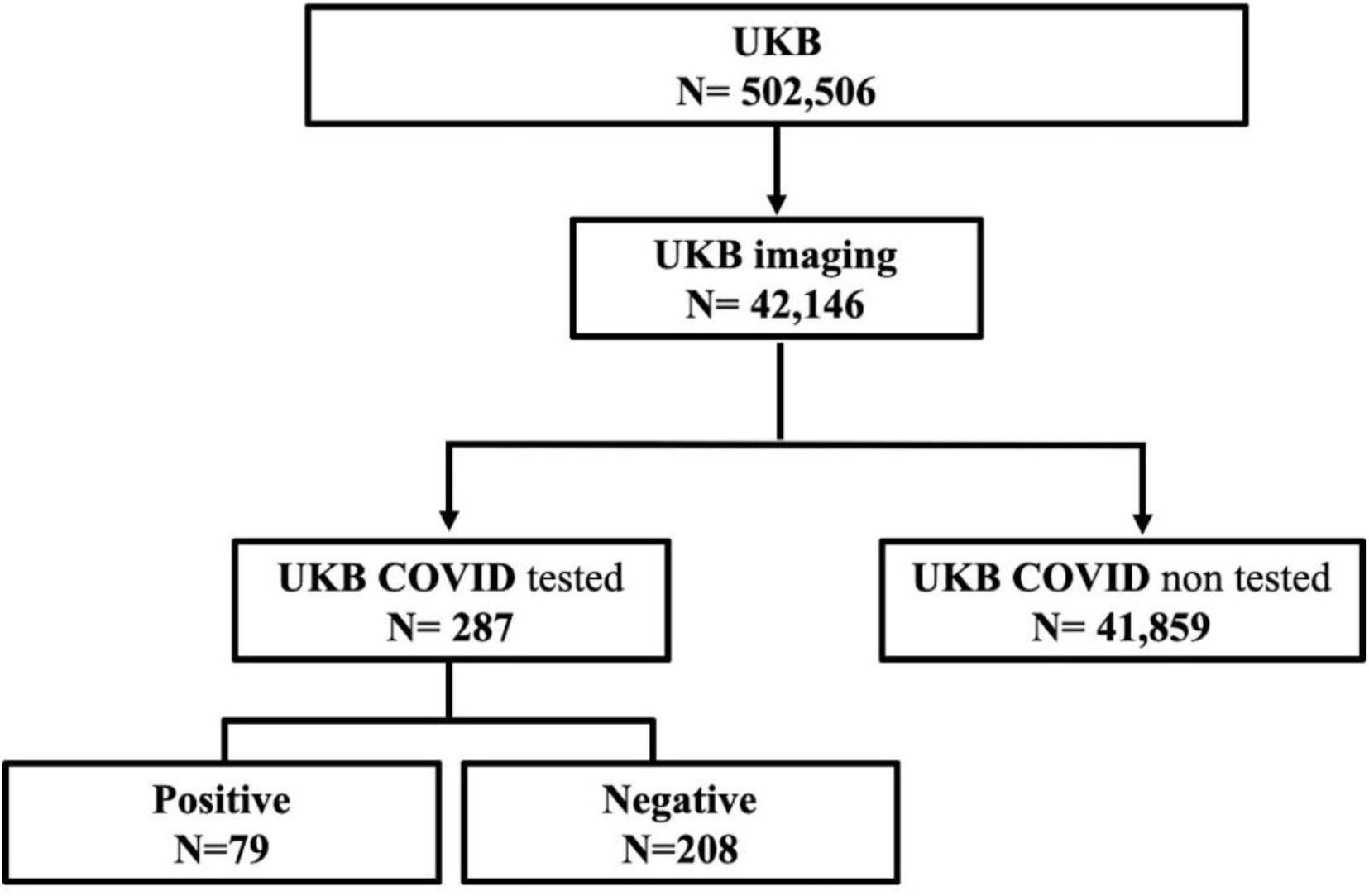
Summary of UKB data available.

Patient meta-data including information on COVID-19 testing status, demographics, diabetes status, cardiometabolic risk factors and genetic variants associated with liver disease were available. No biochemistry data was available for ALT, AST, albumin or GGT levels. MRI measurement of liver fat (proton density fat fraction, PDFF, measured in %) and fibroinflammation (iron corrected T1 mapping, cT1, measured in ms) biomarkers were obtained. Both liver fat and cT1 have been included in the large cohort UK Biobank Study (UKB) (16,17) due to the repeatability and reproducibility of LiverMultiScan (18), and suitability for large-scale population imaging. Liver fat and cT1 are both continuous variables, with population reference ranges and cut-offs for disease previously described. Liver fat > 5% is diagnostic of fatty liver disease (19); liver fat > 10% is considered to qualify as severe fatty liver disease and is a commonly used inclusion criteria in clinical trials of patients with non-alcoholic steatohepatitis (NASH)(20).

A liver cT1 greater than 825ms has been shown to predict liver-related clinical outcomes (14), comparably to histological fibrosis staging.

UKB has released COVID-19 test results from 16 March 2020 onwards, with the majority of SARS-CoV-2 positive tests coming from patients with symptoms in hospital due to the nature of UK testing.

We accessed UKB to determine datasets related to liver health using application 9914 (Determining the Outcomes of People with Liver Disease). The UKB has approval from the North West Multi-Centre Research Ethics Committee (MREC) and obtained written informed consent from all participants prior to the study.

### Statistical analysis

Statistical analysis was performed using R software (version 3.6.1) with a p-value less than 0.05 considered statistically significant. Descriptive statistics were used to summarise baseline participant characteristics. Mean and standard deviation (SD) were used to describe normally distributed-continuous variables, median with interquartile range for non-normally distributed, and frequency and percentage for categorical variables. Mean difference in biomarker values between those who tested positive for COVID-19 and those who did not were compared using the Mann-Whitney-U test, and difference in the counts of the binary outcomes of the self-reported conditions were compared using chi-squared tests. To discriminate patients who tested positive from those who did not, we measured for count data computing the summary measures of risk and a chi-squared test for difference in the observed proportions from count data, presented in a 2 by 2 table with the function *epi.2by2* from the library *epiR* in R. Subsequently, univariable logistic regression analysis was performed for all the potential predictors which included liver fat, cT1, age, sex, ethnicity, body mass index (BMI), self-reported diabetes and self-reported hypertension status. This was performed using the GLM function in R and the brglm2 tool that aims to reduce the bias observed when results are sparse. Risk scores were extracted from the odds ratio estimates of having a positive rest as calculated in the logistic regression model with confidence intervals calculated with the Wald’s test. Pearson’s Chi-square test of independence was used to investigate the relationship of the genetic variation and the result on the COVID-19 test.

## RESULTS

Participant characteristics for each of the groups evaluated in this study are reported in table1. In the symptomatic and tested population, male participants were more likely to test positive for COVID-19. Comparing those that tested positive to the untested population, being male, non-white-British, and diabetic all increased likelihood of being symptomatic and receiving a positive COVID-19 test. The median age of those being symptomatic and testing positive was lower than in the untested population.

**Table 2:**
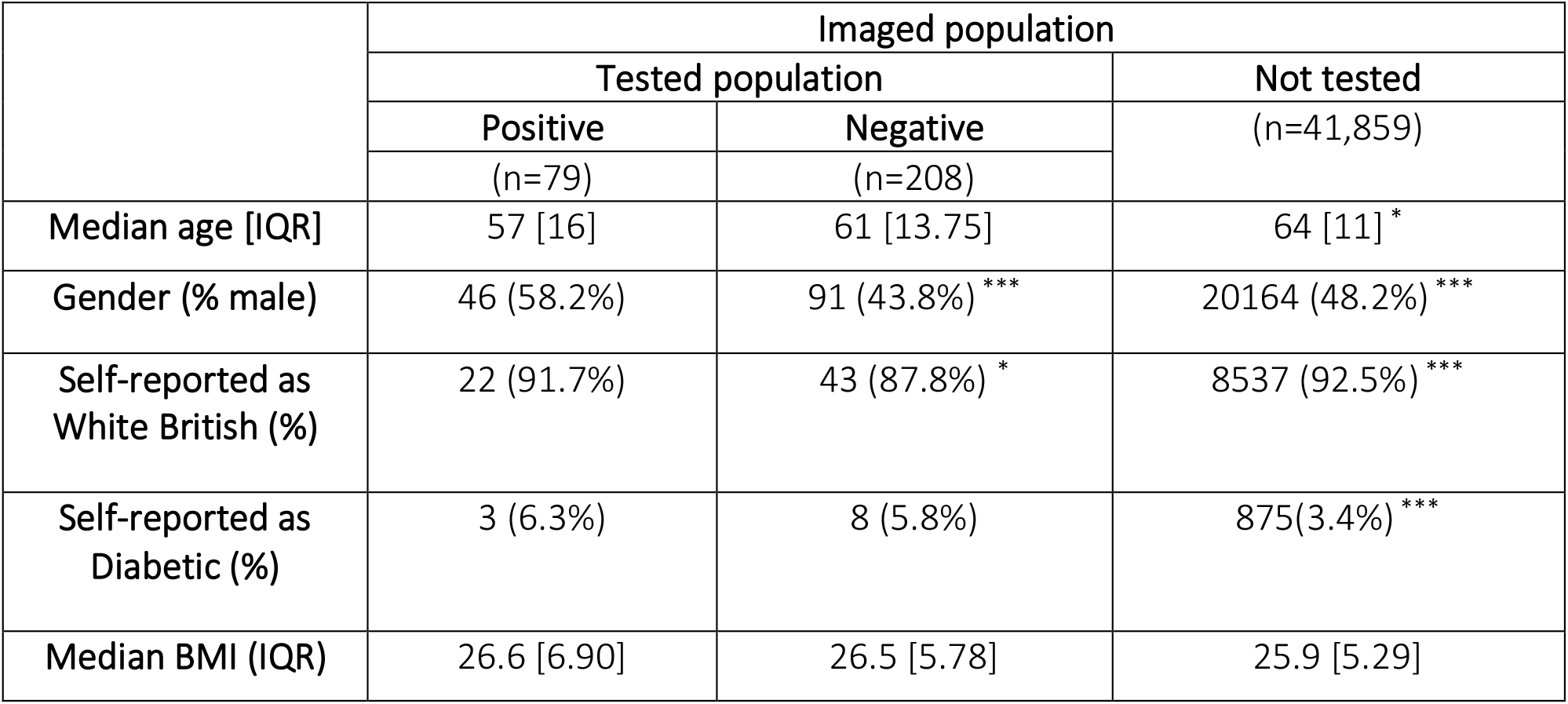
Participant characteristics – Data reported as median [interquartile range] or n (%). Significance of the negative test group versus the positive test group, and the not tested group versus the positive test group are indicated by *p≤0.05, **p≤0,01, ***p≤0.001. Abbreviations: BMI, body mass index; IQR, interquartile range.

|  | Imaged population |  |  |
| --- | --- | --- | --- |
|  | Tested population |  | Not tested<br>(n=41,859) |
|  | Positive | Negative |  |
|  | (n=79) | (n=208) |  |
| Median age [IQR] | 57 [16] | 61 [13.75] | 64 [11] * |
| Gender (% male) | 46 (58.2%) | 91 (43.8%) *** | 20164 (48.2%) *** |
| Self-reported as White British (%) | 22 (91.7%) | 43 (87.8%) * | 8537 (92.5%) *** |
| Self-reported as Diabetic (%) | 3 (6.3%) | 8 (5.8%) | 875(3.4%) *** |
| Median BMI (IQR) | 26.6 [6.90] | 26.5 [5.78] | 25.9 [5.29] |

Table 2 reports the associations between liver fat and fibroinflammation with the likelihood of being symptomatic and testing positive for COVID-19. A normal liver fat (< 5%) is protective against being symptomatic and testing positive, whereas increasing severity of fatty liver disease, or evidence of liver fibroinflammation, increases the likelihood of being symptomatic with COVID-19 (Figure 2).

**Figure 2:**
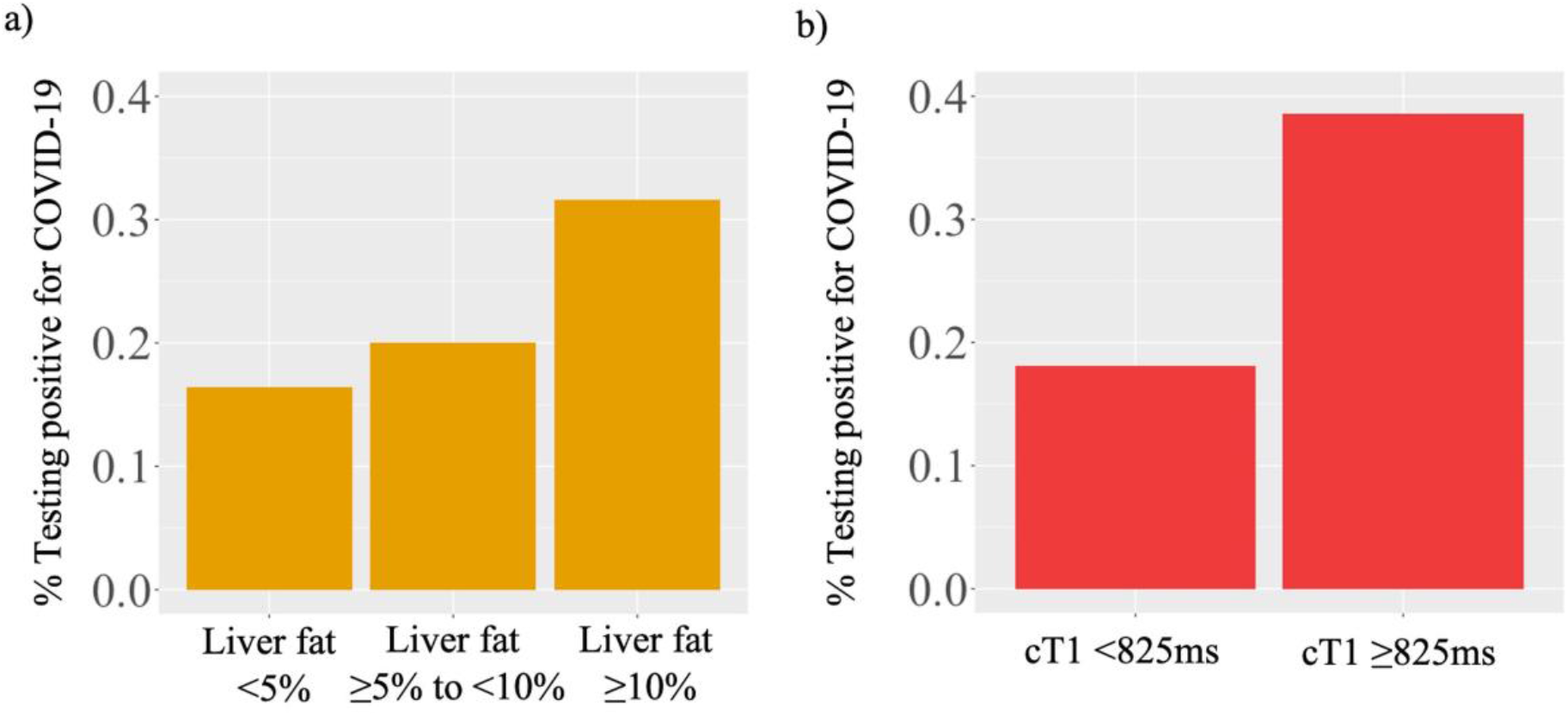
Stepwise increase in percentage of participants being symptomatic and testing positive for COVID-19 with pre-existing fatty liver disease (a) and liver fibroinflammation (b). Abbreviations: COVID-19, Coronavirus disease.

**Table 2:** Associations between imaging biomarkers of liver health and likelihood of being symptomatic and testing positive for COVID-19. Abbreviations: COVID-19, Coronavirus disease; CI, confidence interval. Significance is indicated by *p≤0.05.

|  | Number of participants | Symptomatic and tested for COVID-19 | Positive test for COVID-19 | Odds ratio | Odds ratio 95% CI | p value |
| --- | --- | --- | --- | --- | --- | --- |
| Normal liver fat (<5%) | 30409 | 203 | 50 | 0.66 | (0.42-1.05) | 0.08 |
| Liver fat ≥ 5% | 11737 | 84 | 29 | 1.5 | (0.95-2.38) | 0.08 |
| Liver fat ≥ 10% | 4743 | 39 | 15 | 1.85 | (1.05-3.25) | 0.03* |
| Fibroinflammation (cT1≥825) | 1296 | 14 | 5 | 2.13 | (0.86-5.29) | 0.09 |
| Total | 42146 | 287 | 79 |  |  |  |

Univariate analysis of the imaging biomarkers and known exposures linked to COVID-19 positivity revealed having liver fat ≥10% (P< 0.03), being obese (P< 0.03), being of non-white ethnicity (P< 0.02) and having hypertension(P< 0.04) all significantly increased the odds of testing positive for COVID-19 by 1.85 (CI: 1.05–3.25), 1.87 (CI: 1.06- 3.3), 2.02 (CI: 1.11–3.67) and 2.1 (CI: 1.05–4.21) respectively (figure 3).

**Figure 3:**
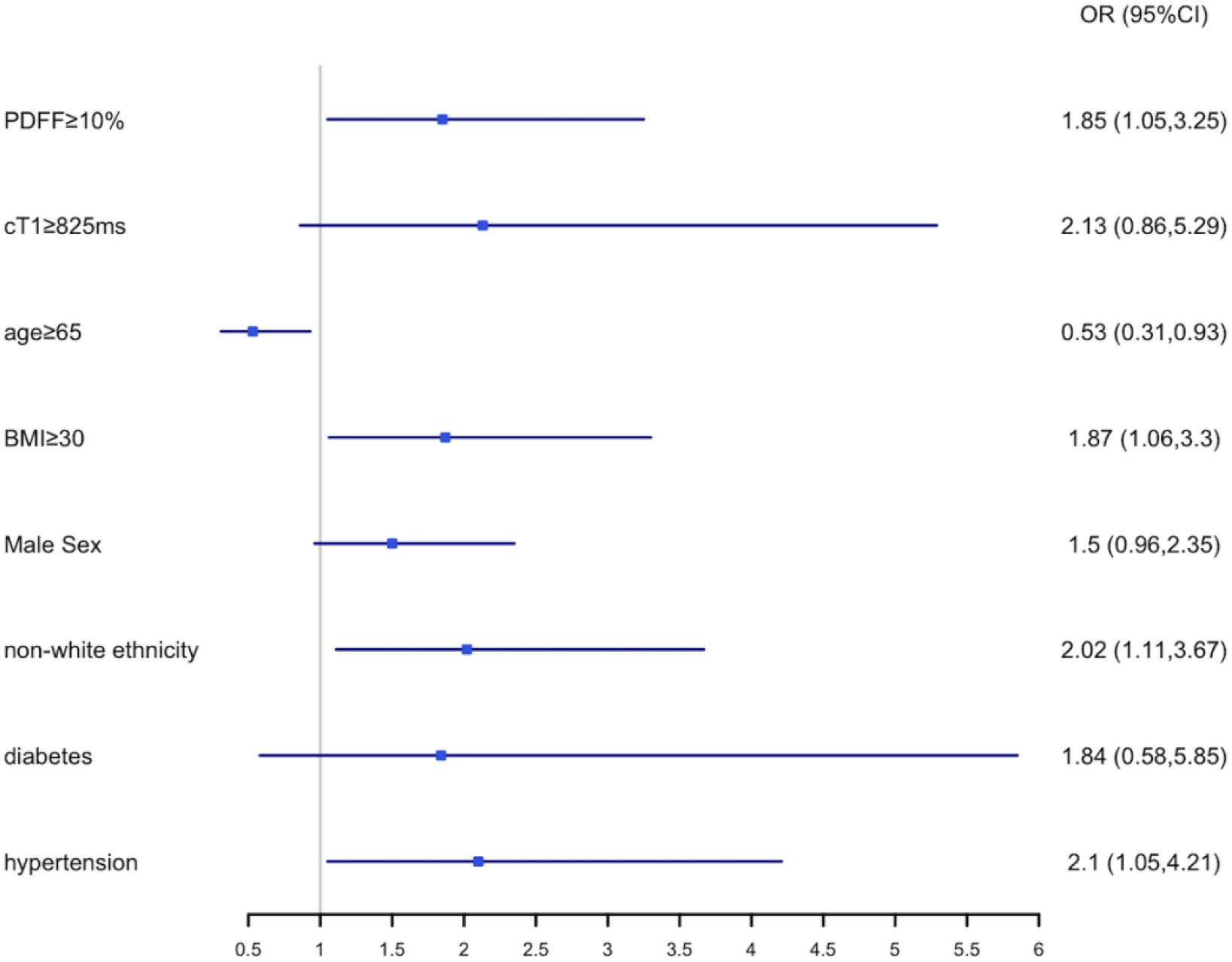
Univariate logistic regression. Comparison of participants who tested positive versus those who do not have test results available or tested negative. Abbreviations: OR, odds ratio; PDFF, proton density fat fraction; BMI, body mass index.

Furthermore, the 32.7% of obese patients with liver fat ≥10% had a higher likelihood of being symptomatic and testing positive for COVID-19 (OR: 2.96, p = 0.02). In contrast, the 37.2% of obese patients with normal liver fat (OR: 0.36, p = 0.09) did not (Table3).

**Table 3:** Associations between obesity and liver fat with the likelihood of being symptomatic and testing positive for COVID-19. Abbreviations: COVID-19, Coronavirus disease; CI, confidence interval. Significance is indicated by *p≤0.05.

|  | Not Obese (BMI < 30 kg/m<sup>2</sup>) |  | Obese (BMI ≥ 30 kg/m<sup>2</sup>) |  |
| --- | --- | --- | --- | --- |
|  | Odds ratio (CI) | P value | Odds ratio (CI) | P value |
| **Normal Liver Fat (<5%)** | 0.93 (0.45-1.96) | 0.86 | 0.36 (0.10 – 1.26) | 0.09 |
| **Moderate fatty liver disease (liver fat 5-9.9%)** | 1.25 (0.55-2.83) | 0.60 | 1.65 (0.37-7.37) | 0.51 |
| **Severe fatty liver disease (liver fat ≥10%)** | 0.69 (0.17-2.86) | 0.61 | 2.96 (1.12- 7.78) | 0.02* |

## DISCUSSION

The aim of this study was to explore the hypothesis that pre-existing liver disease increases the risk of having symptomatic COVID-19. We report on a cohort of 42,146 participants from the UKB with complete liver imaging, of which 287 have COVID-19 test results available with 79 participants testing positive. Our study demonstrates that in addition to the previously-reported risk factors of male gender, non-white-British ethnicity, and obesity (1–3), liver fat is also a significant risk factor for having symptomatic COVID-19, with a person testing positive for COVID-19 being 1.85 times more likely to have pre-existing severe fatty liver disease. To put that into context, 4743 people in this cohort of 42,146 had severe fatty liver disease, suggesting that 11% of the UK population carries this higher risk for COVID-19. Several risk factors have been identified from early studies in China (1), Europe (2) and the US (3) that lead to severe infection, including increased age, male sex, non-Caucasian ethnicity and the presence of pre-existing co-morbidities, such as cardiovascular and metabolic conditions, including diseases of the liver. Unlike previous studies, our research does not find increasing age as a risk factor in this cohort, but this is likely an artefact of the limited age range in the UKB population and increasing likelihood of exposure to the virus in people of working age versus those that had recently retired.

In line with our findings, previous studies have reported BMI as a risk factor for COVID-19, but the mechanism(s) of excess weight in conferring additional risk remain unclear. In this study, more than a third of obese participants (37.2%) had normal liver fat and were not at increased risk of being symptomatic and testing positive for COVID-19. In contrast, obese patients with liver fat ≥10%, representing approximately one third of the obese population, did have significantly higher odds of being symptomatic and testing positive for COVID-19. This highlights the important independent role of liver fat, in addition to obesity, in the severity of, and susceptibility to, COVID-19.

Due to the nature of UK testing for COVID-19 over the period these data were collected, only those symptomatic for the disease to the extent that hospitalisation was warranted will have had test results available. Given the high prevalence of fatty liver disease in the general population, combined with the utility of lifestyle-interventions (22), and new therapeutics that have demonstrated clear reversal of fatty liver disease in weeks, patients with high liver fat may benefit from going on calorie restriction diets and pharmacotherapeutic interventions. Investigational therapies such as NGM282 and Resmetirom have been shown to reduce liver fat in a matter of weeks (23,24), but need to be trialled specifically for utility in this context of use. More generally, public health measures should be considered to protect the 11% of the UK population that has liver fat greater than 10%, as their risk of experiencing COVID-19 with sufficient severity to require hospitalisation has more than doubled.

The main limitation of our study is the relatively small proportion of UKB participants for which testing results are available. This analysis was performed on the second release of COVID-19 testing data and subsequent analyses as more data are released will provide additional power to investigate a larger number of exposures. This limitation is largely a result of UK policy at the time to only test those patients requiring hospitalisation; however, one benefit of this is policy is the ability to infer that those participants who tested positive had severe COVID-19 disease requiring hospitalisation. This inference will not be possible in future releases of COVID-19 testing data following changes in the UK testing policy to support widespread testing.

## CONCLUSIONS

This study highlights the potentially important role of liver disease in COVID-19 severity. As more data becomes available through the UKB, further investigations into the associations of liver health with other demographic characteristics and co-morbidities may reveal more detailed relationships, for example, between multiple risk factors and the development of severe COVID-19. Given the high prevalence of fatty liver disease, and the availability of new treatments specifically for fatty liver disease, these results have the potential to inform public health policy around the management of this at-risk population.

## Data Availability

All source data (including raw and processed MRI data and non-imaging measures) is available from UK Biobank via their standard data access procedure. See the link below.

https://www.ukbiobank.ac.uk/register-apply

## ACKNOWLEDGEMENTS

This research has been conducted using the UK Biobank Resource under application 9914.

